# TMS-induced modulation of brain networks and its associations to rTMS treatment for depression: a concurrent fMRI-EEG-TMS study

**DOI:** 10.1101/2024.12.24.24319609

**Authors:** Hengda He, Xiaoxiao Sun, Jayce Doose, Josef Faller, James R. McIntosh, Golbarg T. Saber, Sarah Huffman, Linbi Hong, Spiro P. Pantazatos, Han Yuan, Lisa M. McTeague, Robin I. Goldman, Truman R. Brown, Mark S. George, Paul Sajda

## Abstract

Transcranial magnetic stimulation (TMS) over the left dorsolateral prefrontal cortex (L-DLPFC) is an established intervention for treatment-resistant depression (TRD), yet the underlying therapeutic mechanisms remain not fully understood. This study employs an integrative approach that combines TMS with concurrent functional magnetic resonance imaging (fMRI) and electroencephalography (EEG), aimed at assessing the acute/immediate effects of TMS on brain network dynamics and their correlation with clinical outcomes. Our study demonstrates that TMS acutely modulates connectivity within vital brain circuits, particularly the cognitive control and default mode networks. We found that the baseline TMS-evoked responses in the cognitive control and limbic networks significantly predicted clinical improvement in patients receiving a novel EEG-synchronized repetitive TMS treatment. Furthermore, this study explored the brain-state dependent effects of TMS, as the brain-state indexed by the phase of EEG prefrontal alpha oscillation. We found that clinical outcomes in this novel treatment are linked to state-specific TMS-modulated functional connectivity within a pivotal brain circuit of the L-DLPFC and the posterior subgenual anterior cingulate cortex within the limbic system. These findings contribute to our understanding of the therapeutic effects underlying TMS treatment in depression and support the potential of assessing state-dependent TMS effects in TMS timing target selection. This study emphasizes the importance of personalized timing of TMS for optimizing target engagement of specific clinically relevant brain circuits. Our results are crucial for future research into the development of personalized neuromodulation therapies for TRD patients.

## Introduction

Transcranial magnetic stimulation (TMS) over the left dorsolateral prefrontal cortex (L-DLPFC) is a Food and Drug Administration-approved treatment for depression. TMS therapy has demonstrated efficacy and safety in the treatment of patients with treatment-resistant depression (TRD)^1,2^. However, the mechanism of action underlying the therapeutic effects of TMS is still unclear. Substantial evidence has shown that TMS not only stimulates the superficial cortex site right underneath the coil, but also has transsynaptic effects on deep brain circuits associated with the stimulation site^3–7^. Assessing such TMS-induced effects on brain circuits allows the quantification of network perturbation and identifies measures relevant to clinical improvement. To investigate TMS-induced effects, the propagation pattern of induced activity from the L-DLPFC to various downstream regions has been established in many studies^8–10^. However, such effects appear to be highly heterogenous both at the stimulation site^11^ and at the associated networks distal to the stimulation site^12,13^, with reports of such stimulation both increasing and decreasing neuronal activity, depending on the region, network, and stimulation parameters. To directly assess TMS-induced acute effects on the brain, recent studies combined TMS with concurrent functional magnetic resonance imaging (fMRI) acquisitions, which allows monitoring of the acute/immediate subsequent effects of the TMS on brain dynamics. For example, Vink et al. investigated the propagation pattern of TMS-induced activity from the L-DLPFC to the subgenual anterior cingulate cortex (sgACC)^13^. Oathes et al. also assessed TMS-evoked response in the sgACC and found its pre-treatment magnitude and the post-treatment changes are both associated with depression improvement^6^. Despite these encouraging results, research on this topic is relatively sparse, and more studies are warranted to examine and quantify the TMS-induced acute effects on brain networks and characterize the variability between patients, which might inform prognosis in depression treatment^14^.

In addition to assessing TMS-evoked response at a particular brain region, it is also important to explore how the induced local activity can drive modulations throughout large-scale brain network systems^15^. Numerous studies have explored the brain circuits affected by TMS perturbation. These efforts include investigating the lasting after-effects on brain connectivity minutes after a TMS session in offline setups, as well as assessing the immediate, acute effects using concurrent TMS-fMRI in online setups^16^. For example, previous studies have shown that TMS applied to the L-DLPFC attenuates hyper-connectivity between sgACC and the default mode network (DMN) in depression patients^17^, and TMS might modulate the abnormal or symptom-related network connectivity in depression^18–20^. These studies provide evidence for the mechanisms underlying its therapeutic effects. However, these TMS-induced effects on brain connectivity were assessed in an offline fashion. It remains unclear whether these lasting after-effects of TMS-induced modulation reflect direct engagement of targeted brain circuits or reflect indirect compensatory effects, such as induced adaptive plasticity or short-term reorganization across brain networks^21,22^. Even though some studies have shown promising results on the consistency of these acute and offline-lasting effects of TMS^23^, more studies are needed to confirm the relationship between online-acute and offline-lasting effects of TMS-induced modulation. Currently, assessing the TMS-induced online-acute modulation with concurrent TMS-fMRI still provides stronger evidence for the direct engagement of target brain circuits compared to the offline setups^22^.

While great progress has been made to integrate TMS with structural and functional MRI on the optimization of TMS spatial targets for depression treatment^24–30^, relatively little has been explored on the optimization of TMS timing for target engagement^31–36^. Based on the substantial evidence of state-dependent effects of TMS on the brain and behavior^22,37–41^, it is reasonable to hypothesize that TMS timing relative to the state of the brain matters for the TMS treatment of depression. However, the definition of brain state varies across subfields and contexts, and brain state fluctuates at different timescales^41,42^. In this study, we derive a brain state index varying at a relatively short timeframe. Specifically, we proposed to use the phase of prefrontal alpha oscillation in the electroencephalography (EEG) signal as an index of brain state. Many studies have demonstrated that the alpha phase is associated with an active inhibitory mechanism, and the timing of sensory stimulation relative to the alpha phase influences perception^43–47^. These studies indicate a gating mechanism of prefrontal alpha oscillation, with distinct phases reflecting different neural excitability, thus gating the information flow across brain networks. Thus, we hypothesized that prefrontal alpha oscillation might gate the propagation of TMS-induced effects across brain networks, and by potentially targeting the TMS pulses to the personalized phase in the alpha cycle, we can induce a stronger effect at the distal target. Here, we integrated EEG with TMS to track brain state, which is used for personalized TMS pulse timing optimization. In our previous studies, we have shown the benefit of EEG-synchronized repetitive TMS (rTMS) treatment, where we observed progressive entrainment effects over the sessions of rTMS treatment^48^, which are related to better TMS antidepressant response^31^. In another study, our group demonstrated that TMS-induced effects in the circuit between DLPFC and sgACC depended on the EEG prefrontal alpha phase^35^. However, TMS-induced modulation of large-scale brain network systems across the whole brain was not assessed, and their longitudinal changes over the pre-and post-treatment scans have not been explored.

Here, we aim to investigate and quantify the TMS-induced acute modulation on brain networks and to test its associations with the clinical improvement in rTMS treatment for depression. As shown in Fig. 1, in this study, the treatment was designed as follows: 1) At baseline, an integrated fMRI-EEG-TMS (fET) instrument was developed and used as a pre-treatment scan to select the personalized TMS timing target (optimum phase), which was defined as the prefrontal alpha phase that produced the largest blood-oxygen-level-dependent (BOLD) signal increase in the dorsal anterior cingulate cortex (dACC); 2) During the rTMS treatment, patients were randomized into two groups with either rTMS pulses synchronized to the personalized optimal phase as the timing target (SYNC group) or delivered at a random phase (UNSYNC group).

**Fig. 1.**
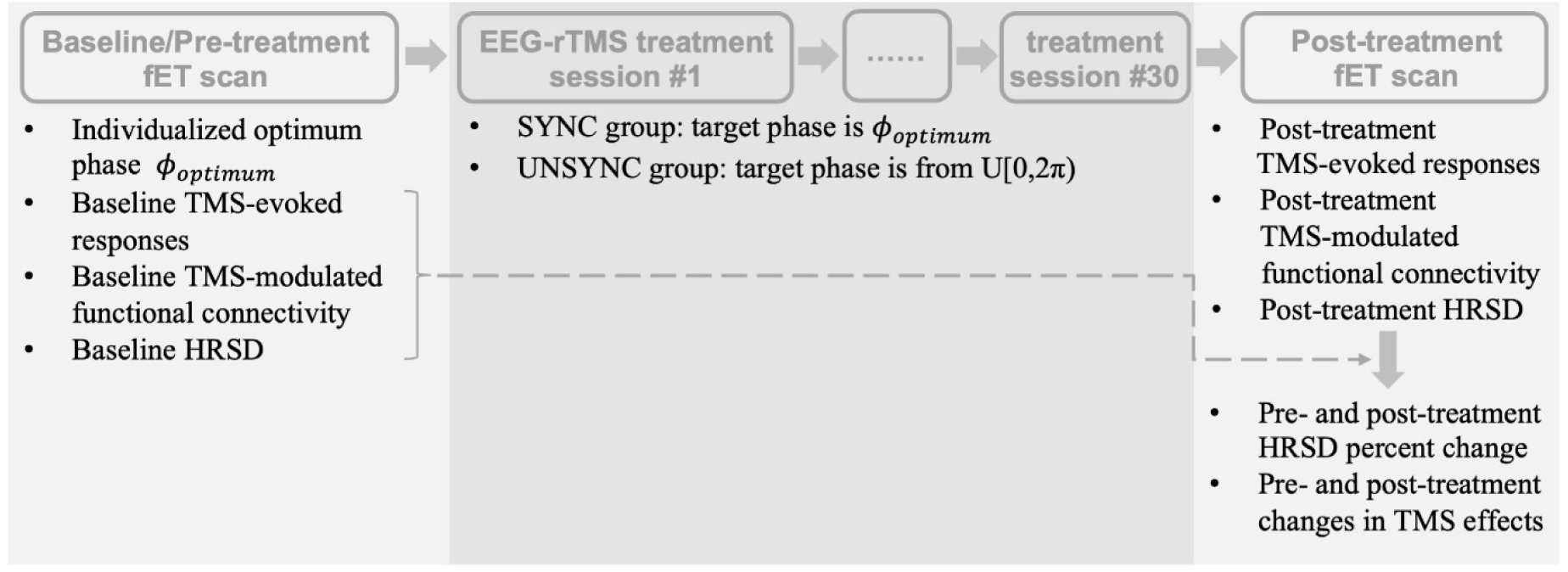
Experimental procedure and data analyses. TMS-induced effects were assessed at both pre-and post-treatment fMRI-EEG-TMS (fET) scans. Both baseline TMS-induced effects and the longitudinal changes in TMS effects were related to the clinical outcome (HRSD percent change). HRSD, Hamilton Rating Scale for Depression.

Patients were treated with 30 rTMS sessions over six weeks; 3) After the treatment, another fET scan was acquired. In this study, we aimed to explore how TMS-induced acute effects propagate through brain network systems. Specifically, we assessed TMS-evoked BOLD responses and connectivity modulations, hypothesizing that TMS over the L-DLPFC would modulate not only the local brain networks but also distal networks associated with depression, such as the cognitive control network (CCN) and the DMN. Furthermore, we tested the state-dependent effects of these modulations using fET scans, with prefrontal alpha oscillation phase indexing brain state. Additionally, we investigated TMS-induced acute effects on brain networks before and after a six-week rTMS treatment. We hypothesized that these TMS-induced effects, both at baseline and post-treatment, would be associated with clinical response. By quantifying the TMS-induced acute effects and state-dependent effects, this study is important for future efforts to temporally optimize TMS targeting in the treatment of depression, with potential implications for personalized treatment strategies in depression^31,32,35,48^.

## Results

Patients in the SYNC group (N = 15) have Hamilton Rating Scale for Depression (HRSD) scores of 30.50 ± 4.35 (mean ± SD; SD, standard deviation) at baseline and HRSD scores of 14.50 ± 7.68 (mean ± SD) post-treatment, with a percent improvement of 52.56% ± 25.37% (mean ± SD). Patients in the UNSYNC group (N = 13) have HRSD scores of 28.40 ± 7.29 (mean ± SD) at baseline and HRSD scores of 13.70 ± 8.22 (mean ± SD) post-treatment, with a percent improvement of 55.31% ± 19.04% (mean ± SD). There is no significant difference in the HRSD percent improvement between groups (p > 0.78). More details on participants recruitment and demographic information are in the “Methods” section.

### TMS-induced acute/immediate effects on whole-brain BOLD signal at baseline fET scan

The group-level TMS-induced BOLD activation map is shown in Fig. 2A (permutation test with FSL Randomise^49^; FWE-corrected p < 0.05). TMS significantly elevated BOLD signals in various brain regions, including dACC, occipital areas, insula, and thalamus, which are consistent with the literature^50^. Next, we sought to computationally quantify these TMS-induced acute effects on brain networks. Specifically, we quantified both the amplitude and spatial extent of TMS-evoked responses across brain networks. These quantifications allow us to assess the engagement of the neural circuits under neuromodulation. We hypothesized that specific neural circuits engaged at the baseline acquisition could predict the following rTMS antidepression response, and any significant results might potentially provide evidence of their mechanistic contribution to the rTMS therapeutic effects^6^. As shown in Fig. 2B, TMS evoked the strongest response in the salience/ventral attention network (subnetwork A, right hemisphere (RH)) and the smallest response in the limbic network (subnetwork B, left hemisphere (LH)), with the highest inter-subject variability in the somatomotor netwosrk (subnetwork A, RH) and the lowest inter-subject variability in the limbic network (subnetwork B, RH). Fig. 2C illustrates the spatial coverage of brain networks under the propagation of TMS-induced effects, where the somatomotor network (subnetwork B, RH) has the highest spatial extent evoked by TMS, and limbic network (subnetwork B, RH) has the smallest spatial extent evoked. As for the spatial extent, we observed the largest and smallest inter-subject variabilities in the visual peripheral network (RH) and limbic network (subnetwork B, RH), respectively.

**Fig. 2.**
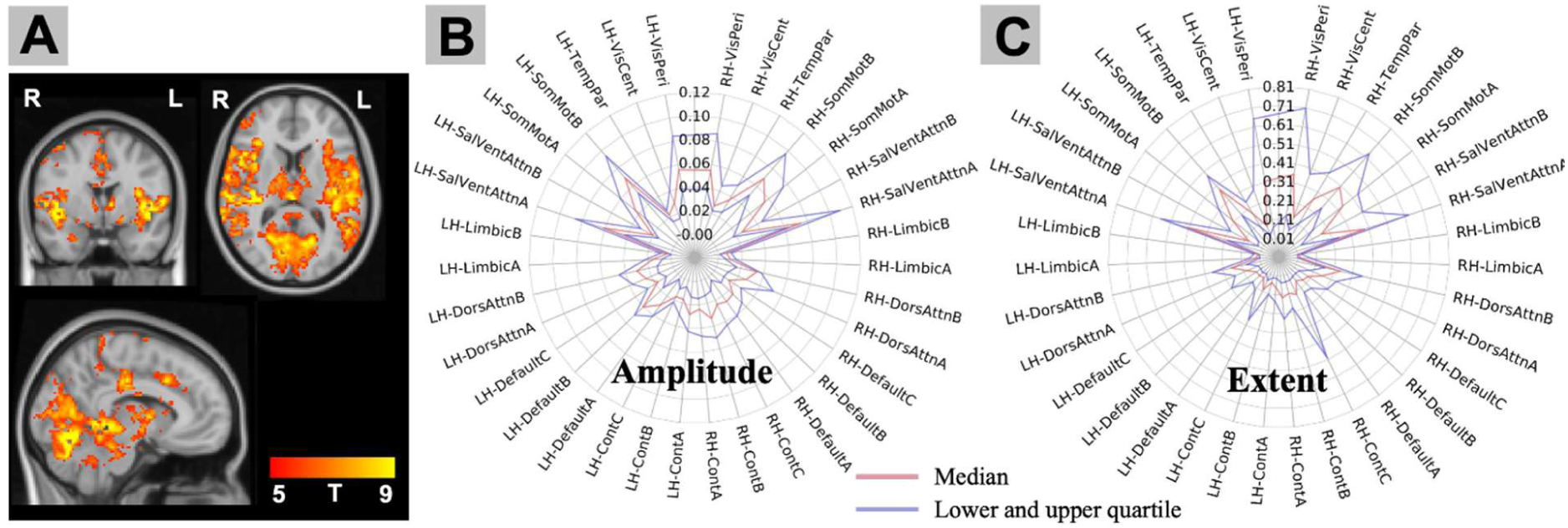
Quantification of baseline TMS-evoked responses on whole-brain BOLD signal. (A) group-level activation map (t-value; p < 0.05 FWE multiple comparison correction; mixed effect); (B) Amplitude of TMS-induced BOLD response in brain networks (Schaefer atlas brain parcellation); (C) The spatial extent of TMS induced-activity propagation coverage through the networks was computed (percentage coverage). The red line indicates the median across subjects. The blue lines indicate the lower and upper quartile across subjects. TMS evoked the strongest response in the SalVentAttn network (subnetwork A, RH), with the highest inter-subject variability in the SomMot network (subnetwork A, RH). We found that the SomMot network (subnetwork B, RH) had the highest extent of propagation coverage of TMS-induced activity, and the VisPeri network in the RH had the highest inter-subject variability in the propagation coverage. LH, left hemisphere; RH, right hemisphere; SomMot, somatomotor network; VisPeri, visual peripheral network; SalVentAttn, salience/ventral attention network. Default, default mode network; VisCent, visual central network; Cont, control network; TempPar, temporal parietal network; DorsAttn, dorsal attention network.

### TMS-induced acute/immediate modulation on brain connectivity at baseline fET scan

To examine the TMS-induced acute/immediate modulation of functional connectivity (FC), we performed a whole-brain psychophysiological interaction (PPI) analysis^51–53^. Significant TMS modulations between networks are shown in Fig. 3A (FDR-corrected p < 0.05), with the strongest negative effect on the connectivity between default mode network (subnetwork A) in the LH and default mode network (subnetwork B) in the RH. Of note is that these are all negative effects, and none of the connections showed significant positive effects after multiple comparison corrections. To identify brain regions that are important for potentially facilitating the propagation of TMS-induced effects over networks, we performed hub analysis, where positive and negative node strength was computed by summing across all positive or negative connections associated with each network node, respectively. As shown in Fig. 3B and 3C, the results showed that the positive hubs are mostly within the visual and somatomotor networks, and the negative hubs are regions in the default, control, and salience ventral attention networks. The summarized hub strength results showed that nodes in the somatomotor network (subnetwork B, LH) and salience/ventral attention network (subnetwork A, RH) have the most positive and negative node strength, respectively.

**Fig. 3.**
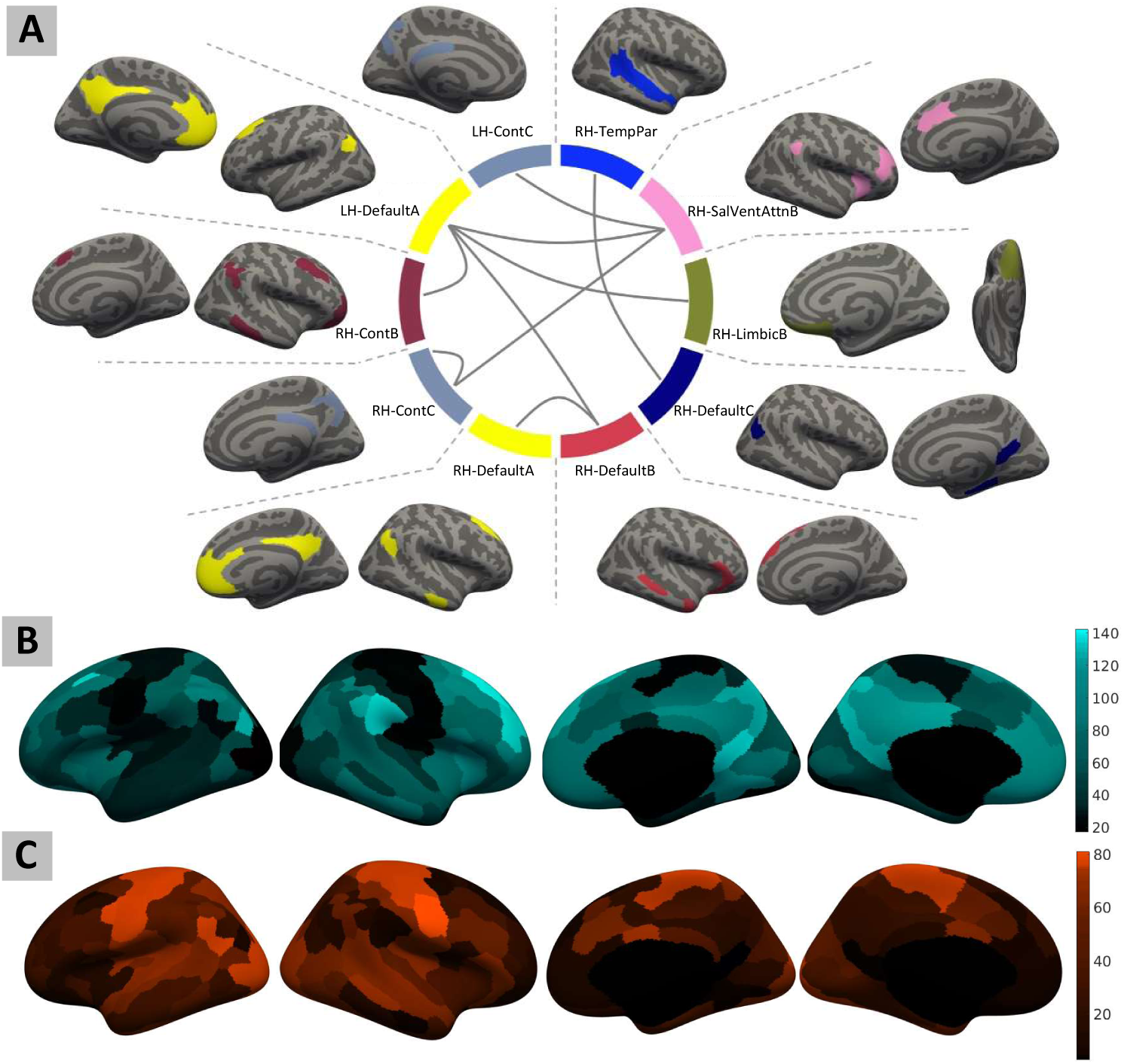
Quantification of TMS-induced functional connectivity at baseline fMRI-EEG-TMS (fET) scan. (A) Group level whole-brain psychophysiological interaction analysis results. TMS induced significant negative effects on the connectivity between cortical networks (FDR multiple comparison corrected p < 0.05). No significant positive effect was observed after multiple comparison correction. (B) and (C) represent negative and positive node strength by computing the sum of all the negative and positive connection weights between one node and all other nodes, respectively. The positive hubs are mostly within the visual and somatomotor networks, and the negative hubs are regions in the default, control, and salience ventral attention networks. LH, left hemisphere; RH, right hemisphere; SalVentAttn, salience/ventral attention network. Default, default mode network; Cont, control network; TempPar, temporal parietal network.

### State-dependency of TMS-induced acute effects and modulation at baseline fET scan

In this section, we investigated TMS state-dependent effects, where we grouped the TMS trials into four phase bins based on their timing relative to the phase of the prefrontal alpha oscillation, resulting in four phase-bin conditions. Then, we identified two TMS trial conditions for each subject: 1) high-load-phase (HLP) condition was defined as the condition where a high TMS evoked response was introduced at the stimulation site (L-DLPFC); 2) low-load-phase (LLP) condition was defined with a low evoked response at L-DLPFC. We assessed the contrast between HLP and LLP conditions, where the whole-brain general linear modeling (GLM) analysis identified regions in the lateral frontoparietal network^54^ as significant clusters (p < 0.001), including bilateral DLPFC and inferior parietal lobule (Fig. 4A). Because HLP and LLP conditions were defined on the BOLD signal only from the L-DLPFC region, activation pattern of their contrast reflects brain areas associated with a higher load of TMS-induced effects on the L-DLPFC, potentially suggesting the whole-brain spreading pattern of TMS-induced response. Then, we hypothesized that this TMS-effects spreading pattern follows brain connectivity. To test this, we compared the spatial pattern of this activation contrast to the seed-based functional connectivity map of the L-DLPFC stimulation site. With different thresholds on the connectivity map, it showed the highest overlap of 28.66% (Dice similarity coefficient (DSC)) with the TMS response contrast map at the group level (Fig. 4B). We replicated these results using the Schaefer atlas L-DLPFC regions of interest (ROIs) near the stimulation site, by also examining HLP and LLP contrast map and seed-based functional connectivity map of each ROI. The results showed a high overlap between the spatial spread of state-dependent effects from L-DLPFC and the connectivity pattern of the L-DLPFC (L-DLPFC in the default mode subnetwork-A: DSC = 25.97%; L-DLPFC in the default mode subnetwork-B: DSC = 37.08%; L-DLPFC in the salience/ventral attention subnetwork-B: DSC = 30.53%; see supplementary figures for details). These results are consistent with the hypothesis that the spatial spread of TMS-induced acute/immediate effects is related to the functional connectivity pattern of the stimulation site^12^.

**Fig. 4.**
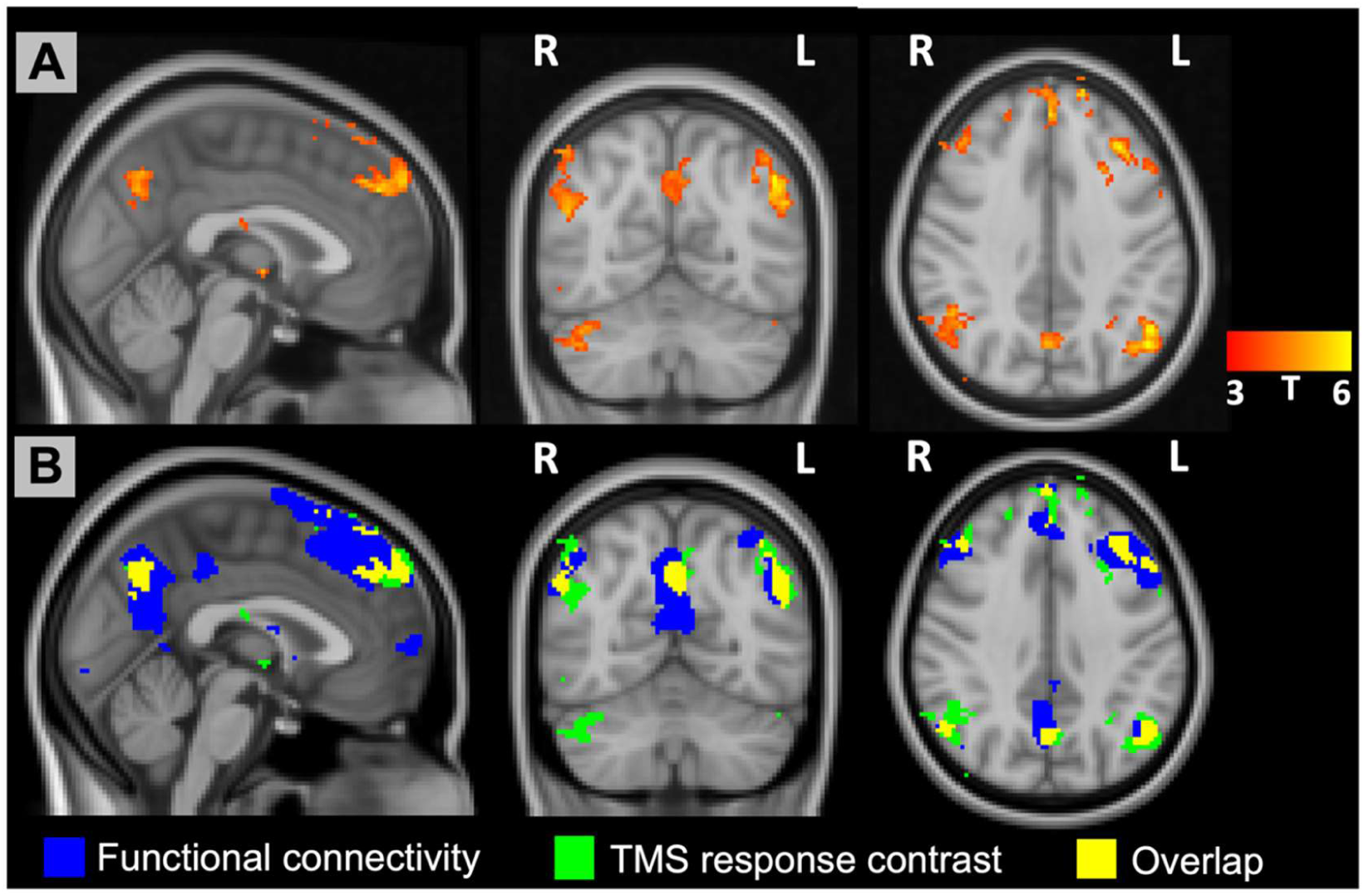
State-dependency analysis of TMS-evoked BOLD response using baseline fMRI-EEG-TMS (fET) scan. (A) TMS response contrast between the conditions of TMS trials in the high-load-phase (HLP) bins and low-load-phase (LLP) bins. Regions in the lateral frontoparietal network were identified as significant clusters (t-value; p < 0.001). Because HLP and LLP conditions were defined based solely on the BOLD signal from L-DLPFC region, the activation pattern resulting from their contrast highlight brain areas associated with greater TMS-induced effects on the L-DLPFC, indicating a whole-brain spreading pattern of the phase-dependent TMS-induced response. (B) Spatial overlap between the TMS response contrast and L-DLPFC seed-based functional connectivity. The TMS response contrast (green) and overlap (yellow) areas encompass the same regions shown in the panel (A). L-DLPFC functional connectivity map showed a network overlapped with the TMS response contrast map, suggesting the propagation of TMS-induced acute effects is related to the functional connectivity.

We also examined whether TMS modulates the connectivity between brain networks differently when the pulses were delivered at different prefrontal alpha phases. Specifically, we used PPI analysis to assess connectivity modulations of the TMS trials in the HLP and LLP bins of the L-DLPFC stimulation site. Our results showed that only the TMS trials in the HLP bins induced significant (FDR-corrected p < 0.05) negative functional connectivity modulations between 1) default mode subnetwork-A in the RH and default mode subnetwork-B in the RH; 2) default mode network (subnetwork A, RH) and subnetwork-A of the control network (RH); 3) default mode network (subnetwork A, RH) and subnetwork-B of the control network (RH). Whereas, no significant functional connectivity modulations were observed for the TMS trials in the LLP bins of the stimulation site. These findings suggest state-specific TMS modulations on the connectivity, rendering the importance of TMS timing optimization to induce stronger modulations on the target brain circuits. We also examined these state-specific modulations for the HLP bins of the Schaefer atlas L-DLPFC ROIs, where no significant modulations were found after multiple comparison correction (see supplementary text for details).

### Associations of TMS-induced acute/immediate effects to the clinical response in rTMS treatment for TRD patients

In this section, we first tested the associations between the quantification of TMS-induced acute/immediate effects at baseline and the clinical response during rTMS treatment. As for the TMS-evoked BOLD response, we observed a significant correlation between the clinical improvement (percent decrease/improvement in the HRSD) and the amplitude of evoked response in the subnetwork B of the control network (LH: r = 0.8591, p < 0.0007; RH: r = 0.8601, p < 0.0007), and limbic network (subnetwork B, RH) (r = 0.8991, p < 0.0002) for the patients in the SYNC group. There is no significant association between the clinical improvement and the amplitude of evoked response in any network for the patients in the UNSYNC group (significance defined as p < 0.05 with Bonferroni correction). We also examined whether the spatial extents of evoked response at baseline pre-treatment scan are associated with clinical improvement, and no significant association was found.

Next, we assessed the longitudinal changes in the TMS-induced acute/immediate effects and tested their associations with the clinical outcome during rTMS treatment. We did not observe any significant longitudinal changes in the TMS-induced acute/immediate effects for each group. However, by pooling the patients from both groups, we found that TMS-evoked BOLD responses in the salience/ventral attention network (subnetwork A) significantly decreased from pre-treatment to post-treatment scan (LH: t = -4.0129, p < 0.0074; RH: t = -4.0403, p < 0.0070). But these longitudinal changes were not significantly associated with the clinical outcome (LH: r = -0.2505, p > 0.4851; RH: r = -0.2662, p > 0.4571).

Finally, we asked if any state-specific TMS modulations on the connectivity at baseline or its longitudinal changes are associated with the clinical outcome during rTMS treatment. We assessed the state-specific effects with the HLP condition. Here, as shown in Fig. 5B, we examined four L-DLPFC ROIs, because the results in our state-dependency analyses suggest that the state-specific effects are sensitive to the spatial specificity of the L-DLPFC ROIs (see supplementary text for details). Specifically, we assessed the connectivity modulated by the TMS trials that evoked the largest response (defined as the HLP condition) at 1) L-DLPFC of EEG F3; 2) L-DLPFC in the subnetwork A of the DMN; 3) L-DLPFC in the subnetwork B of the DMN; 4) L-DLPFC in the subnetwork B of the salience/ventral attention network. This analysis allows us to explore how stronger induced local activities near the stimulation site can drive specific modulations throughout brain networks. Any association between these state-specific connectivity modulations and the clinical outcome might indicate these brain circuits’ involvement in treating TRD patients. We did not observe any significant association between the clinical outcome and the baseline state-specific TMS modulations on the connectivity. Then, we tested the associations between the clinical outcome and the longitudinal changes in the state-specific TMS modulations on the connectivity. As for the state-specific effects associated with the L-DLPFC of the default mode network (subnetwork A), we observed that the clinical outcome is significantly correlated to the changes in the connectivity between L-DLPFC in the default mode network (subnetwork B) and orbitofrontal-cortex in the limbic network (subnetwork B, RH) only in the SYNC group (SYNC group: r = 0.9916, p < 1.4971e-6, FDR-corrected p < 0.01; UNSYNC group: r = -0.3841, p > 0.45). This relationship is still significant after regressing out the baseline connectivity measurement from the longitudinal changes (r = 0.7269, p < 0.0411). However, when testing on the other L-DLPFC regions, we did not observe any significant relationship between the clinical outcome and the connectivity changes after multiple comparison correction. For the connectivity between L-DLPFC and RH orbitofrontal-cortex in the SYNC group, we did observe a trend for the state-specific effects associated with the L-DLPFC in the default mode network (subnetwork B; p < 0.0023), but not L-DLPFC of EEG F3 (p > 0.4730) and L-DLPFC in the salience/ventral attention network (subnetwork B; p > 0.0903). Our results suggest EEG-synchronized rTMS treatment induces functional connectivity changes in specific neural circuits that are associated with the clinical outcome, i.e., state-dependent response in the L-DLPFC (default mode network (subnetwork A)) and the connectivity between L-DLPFC (default mode network (subnetwork B)) and orbitofrontal-cortex (RH, limbic network (subnetwork B)). These results provide insight into the therapeutic effect of rTMS and may inform the design of future rTMS interventions in depression.

**Fig. 5.**
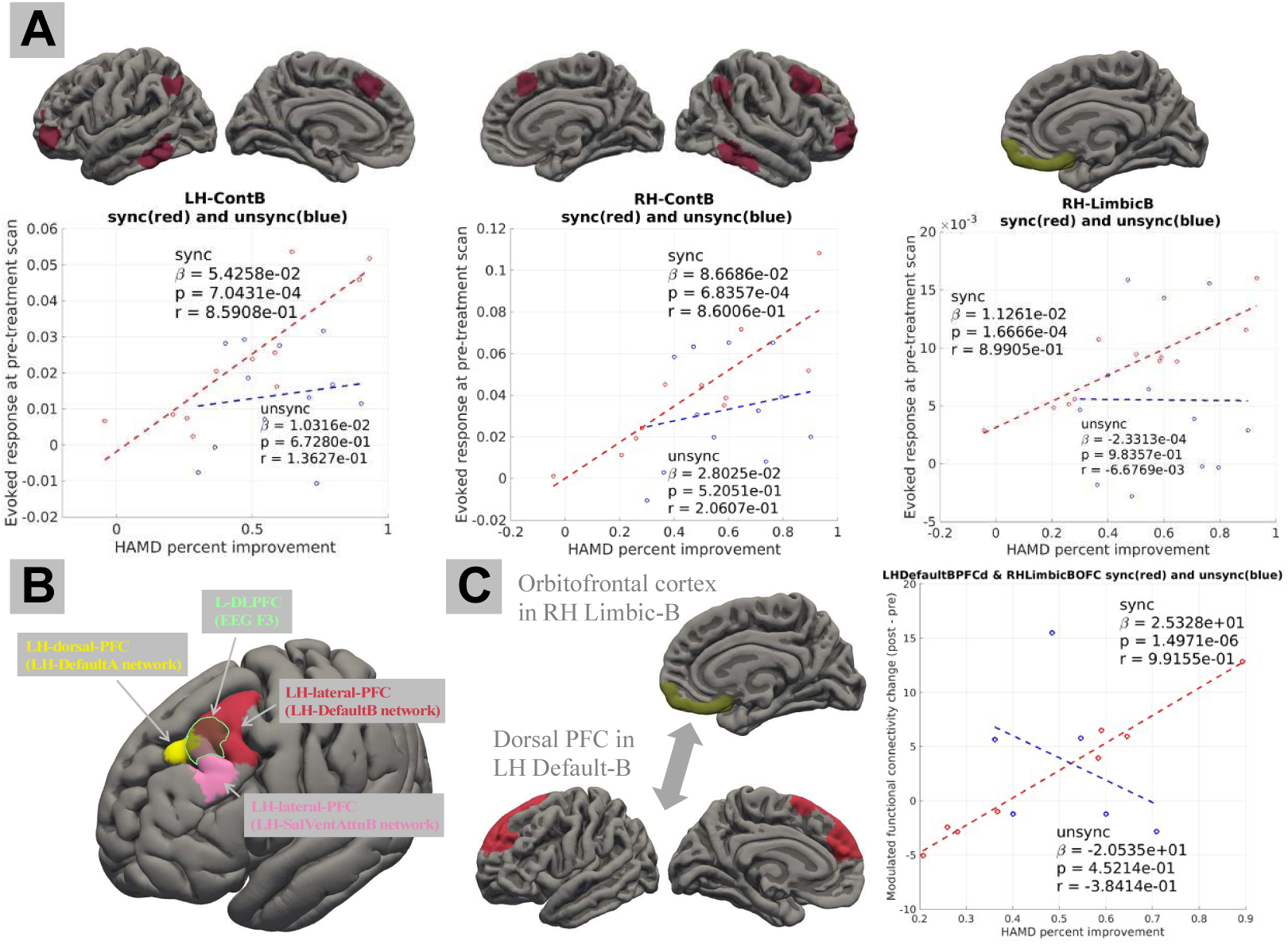
Associations of TMS-induced acute/immediate effects to the clinical response in rTMS treatment for depression patients. (A) We observed a significant correlation between the clinical outcome (percent change in the HRSD) and the pre-treatment TMS evoked response in the bilateral ContB network and RH-LimbicB network only for the SYNC group. (B) State-specific TMS modulations on the connectivity were assessed for four L-DLPFC regions (EEG F3, L-DLPFC in DefaultA, L-DLPFC in DefaultB, and L-DLPFC in SalVentAttnB). (C) As for the L-DLPFC in DefaultA, the pre-and post-treatment state-specific TMS-modulated connectivity changes (between L-DLPFC in the DefaultB network and RH orbitofrontal cortex in LimbicB network) are significantly associated with the clinical outcome only for the SYNC group. No significant result was found for the other three L-DLPFC regions after multiple comparison correction. Panel (A) includes the results of 23 patients (11 SYNC patients and 12 UNSYNC patients) with pre-treatment fET and HRSD available. Panel (C) includes the results of 14 patients (8 SYNC patients and 6 UNSYNC patients) with complete pre-and post-treatment data. HRSD: Hamilton Rating Scale for Depression; fET: integrated fMRI-EEG-TMS. LH, left hemisphere; RH, right hemisphere; SalVentAttnB, salience/ventral attention network (subnetwork B); DefaultA/B, default mode network (subnetwork A/B); ContB, control network (subnetwork B); LimbicB, limbic network (subnetwork B).

## Discussion

In this paper, we investigated the TMS-induced acute/immediate effects on brain networks. Firstly, we quantified its evoked BOLD response and modulated functional connectivity. Then, we tested associations between TMS-induced effects and the clinical outcome. Additionally, we examined whether these TMS-induced acute effects depend on the prefrontal alpha oscillation phase. Our results showed that the spatial spread of TMS-induced phase-dependent effects follows a similar pattern to the functional connectivity of the L-DLPFC stimulation site. When assessing the TMS modulated functional connectivity with trials in the HLP bins (i.e., TMS trials that evoked a higher response at the L-DLPFC stimulation site), we observed significant negative modulations on the control network (subnetwork A and B, RH) and default mode network (subnetwork A and B, RH). No significant modulation was found for trials where TMS was delivered at the LLP timing, and no significant modulation was observed for this state-specific effect associated with other L-DLPFC ROIs near the stimulation site. Finally, we tested the associations between the quantification of TMS-induced acute/immediate effects and the clinical outcome, where TRD patients received a randomized six-week rTMS treatment. We found that the baseline TMS-evoked responses in the bilateral control network (subnetwork B) and limbic network (subnetwork B, RH) significantly predict the following clinical improvement, but only in the group of patients receiving the EEG-synchronized rTMS treatment (SYNC group). Finally, our results showed that the longitudinal changes in the state-specific TMS modulations on the connectivity between L-DLPFC (part of the subnetwork B of the DMN) and right hemisphere orbitofrontal cortex (part of the subnetwork B of the limbic network) was significantly associated with the clinical improvement only for the patients in the SYNC group. The present study assessed TMS-induced acute/immediate effects on brain networks with concurrent TMS-fMRI and investigated the brain-state dependency of the induced effects with simultaneous EEG recordings. The results of its association with clinical outcomes have potential implications for developing efficient and personalized treatments for TRD patients.

The concurrent acquisition of fMRI data during single-pulse TMS enables the investigation of the immediate BOLD response to TMS. This TMS-evoked BOLD response, frequently characterized in recent concurrent TMS-fMRI studies, serves to demonstrate target engagement or its associations with the clinical outcome. For example, Oathes et al. explored the TMS-evoked response in subcortical brain areas and the associated networks, such as the amygdala and sgACC^55^, demonstrating engagements of these circuits underlying neuromodulation. Similarly, Vink et al. assessed the spatial propagation of TMS-evoked responses in the sgACC and across the whole brain^13^. In support of the findings in other literature, these studies highlighted the substantial individual variability in TMS-evoked responses. The observed variability may be attributable to neural activity at the stimulation site^11^ and/or its functional connectivity with specific brain networks, such as the salience network^12^. These observations underscore the importance of characterizing such individual variabilities. Thus, in this study, we proposed to quantify both the amplitude and the spatial propagation extent of the immediate acute TMS-evoked responses. Such quantifications potentially validate target engagement and could have important clinical implications. For instance, a recent study linked the TMS-evoked response in the sgACC to clinical outcomes following rTMS treatment^6^. Their results suggest that a stronger evoked response in the sgACC during the pre-treatment TMS-fMRI session demonstrated better target engagement and subsequently correlated with improved therapeutic outcomes in the treatment. Our findings align with these studies, where we demonstrated that the evoked response in the right orbitofrontal cortex (containing the posterior sgACC) within the limbic network (subnetwork B) during the pre-treatment session is relevant to the clinical outcomes of rTMS treatment. Additionally, we observed that baseline-evoked responses in the bilateral control network (subnetwork B) were significantly associated with clinical outcomes. However, these associations are only significant for the patients in the SYNC group. This suggests that patients exhibiting the strongest evoked responses in the limbic network (subnetwork B, RH) and bilateral control network (subnetwork B) networks at baseline, rendering a better target engagement of these networks, benefit most from EEG-synchronized rTMS treatment. Conversely, the nonsignificant results for patients in the UNSYNC group might imply that the therapeutic effects of unsynchronized rTMS treatment might not be specific to certain brain networks. Moreover, with the longitudinal data, we showed a significant decrease in the evoked response within the salience network after rTMS treatment, which does not correlate significantly with the clinical outcomes. These findings might suggest the potential impact of rTMS on anxiety symptoms, given the established link between the salience network and anxiety ^56,57^. However, further research is warranted to validate this with a larger sample size.

Numerous studies have illustrated that TMS not only influences the stimulation site and its directly connected network but also has effects that propagate to other brain networks^58^. Considering these distributed brain regions collectively within the framework of brain circuits and networks, especially in relation to the impacts of TMS or the characterization of depression patients^59,60^, could enhance our understanding of the therapeutic effects of TMS in depression, and also in the target selection. In support of this literature, we propose to characterize and quantify such effects at the level of network connectivity. Recent studies have demonstrated that rTMS may down-regulate and normalize hyperconnectivity of certain brain networks, such as the DMN^17,61^ and the salience network^62^. Consistent with these findings, our study also reveals that TMS induces significant negative modulations in the connectivity between brain networks, including the DMN, control, and salience networks. These results are consistent with the literature, where Chen et al. observed significant negative TMS modulations between the CCN (lateral frontoparietal network^54^ or central executive network) and the DMN when single-pulse excitatory TMS delivered to the node of CCN^7^. Our results also are consistent with the findings that TMS can normalize depression-related hyperconnectivity associated with DMN^17^. These potentially suggest the engagement of these brain networks in the therapeutic mechanisms of rTMS by normalizing the network hyperconnectivity in depression patients. However, unlike these studies, our concurrent acquisition allows us to assess the acute modulatory effects of TMS on network connectivity and potentially provides more causal insights into the interactions between brain networks. Furthermore, our findings are in line with concurrent TMS-fMRI research, showing that TMS application to the central executive network (CEN) node induces causal negative downstream effects on the DMN^7^. We also found that the negative hubs in the TMS-induced acute modulations on connectivity are located within the default, control, and salience ventral attention networks, while positive hubs are primarily within the visual and somatomotor networks. This reveals a differentiation in the top-down pathways involved in the propagation of TMS-induced acute effects across brain networks, which aligns with the categorized hub types hypothesis^63^. Notably, some studies have reported that the TMS effects on the connectivity of the visual networks contribute to therapeutic outcomes^20,29^. In future studies, it is worth further investigating TMS modulations on visual networks in comparison to its negative modulations on the DMN and cognitive control networks. Thus, further exploration of the excitatory and inhibitory effects of single-pulse TMS at the stimulation site^11^, along with the propagation of TMS effects through both positive and negative connections across distinct brain hub regions, is warranted.

In this study, we utilized the phase of EEG prefrontal alpha oscillation as an indicator of brain state and investigated the brain-state-dependent effects of TMS-induced brain activity. Our results suggest that the propagation of TMS-induced BOLD activity from the L-DLPFC (EEG F3 stimulation site) to regions within the L-FPN is dependent on the EEG prefrontal alpha phase. Specifically, in conditions where TMS trials elicited a stronger response at the L-DLPFC (HLP timing), areas within the L-FPN also exhibited a stronger response to these TMS pulses, suggesting that the state-dependent spread patterns of TMS-induced activity closely follow the functional connectivity of the L-DLPFC, with high spatial similarity. However, when examining other ROIs proximate to the L-DLPFC F3 stimulation site, the results suggest distinct spread patterns, underscoring the spatial specificity inherent to the localization or characterization of the L-DLPFC ROIs. These findings align with existing literature, indicating that the spread of TMS-induced activity from the L-DLPFC stimulation site follows the functional connectivity pattern of the site^12^. Nevertheless, Hawco et al. also reported that this relationship is mediated by the characteristics of the L-DLPFC, particularly depending on its functional connectivity to the salience network. Our results, along with the literature, highlight the critical role of leveraging functional connectivity to guide TMS targeting^24^. However, it should be noted that we assessed the functional connectivity of the L-DLPFC with TMS-evoked responses regressed out to control the confound of TMS-related activations. Future studies should explore the spread of TMS-induced response and functional connectivity based on separate resting-state fMRI sessions to completely rule out the possible interactions between spontaneous brain activity and TMS. In this study, we used the prefrontal alpha oscillation phase as an index of brain state and assessed state-dependent effects on both evoked response and modulated functional connectivity. This choice was motivated by literature evidencing the gating mechanism of the alpha oscillation phase. However, to further elucidate the state dependency of TMS-induced activity, future research should consider other brain state indices, such as the power of alpha or the phase of theta oscillation. In our previous investigations, we demonstrated that the TMS-evoked BOLD response in the cingulate gyrus is dependent on the EEG prefrontal alpha phase^35^. Additionally, we have reported that alpha phase-synchronized rTMS treatment facilitates phase entrainment in patients^48^, with increased entrainment over time correlating with improved rTMS treatment outcome^31^. Altogether, these findings contribute to the development of biomarkers for tracking the efficacy of EEG-synchronized rTMS treatment^64^. Following our previous findings, the current study focused on quantifying the acute effects of TMS on fMRI signals and evaluating its clinical relevance. These results add new perspectives to our understanding of TMS-induced effects on brain networks and also shed light on the therapeutic effects of rTMS treatment in depression.

The observed association between changes in state-specific connectivity modulation and clinical outcomes suggested that patients who achieved a more increased modulated connectivity between the left dorsal-prefrontal cortex and the right orbitofrontal cortex benefitted the most from EEG-synchronized rTMS treatment. This brain circuit, which showed relevance to the clinical outcome, is associated with the personalized prefrontal alpha phase (HLP condition) in which TMS evoked the strongest response in the L-DPFC within the default mode (subnetwork A). Furthermore, to examine spatial specificity, we assessed this relationship with other L-DLPFC ROIs. Our results revealed that, when evaluating the phase-dependent response, the strongest association to the clinical outcome was observed for the L-DLPFC within the subnetwork A of the DMN, compared to the ROIs in the EEG F3 area, the subnetwork B of the DMN and salience/ventral attention network (subnetwork B). However, caution should be taken when evaluating the correlation coefficient results with a small sample size in this study. No such association was observed when using patients in the UNSYNC group as a control group. These findings highlight the potential systematic therapeutic effect of EEG-synchronized rTMS treatment in engaging more specific brain circuits, which might be more directly related to the clinical therapeutic effects in depression, as opposed to the possible widespread effects in the unsynchronized rTMS treatment. The latter may induce effects across a wide range of brain circuits, with TMS pulses during various brain states affecting distinct circuits. Randomly fired TMS pulses at different prefrontal alpha phases could modulate different network connections, with some pulses occurring at the right time actually having beneficial effects. Our findings align with existing literature that the therapeutic mechanism of rTMS treatment is likely to involve the circuit of the right orbitofrontal cortex (containing posterior sgACC) and L-DLPFC^24,65^. Future research with a larger cohort is necessary to validate these results.

This study is subject to several limitations that warrant consideration in the future. Firstly, while our findings are promising, the current study is constrained by a relatively small sample size. Our results demonstrated that quantified effects of TMS are correlated with clinical outcomes only in the SYNC group, and results from the UNSYNC group served as a control. However, a direct comparison of the UNSYNC group’s results with those of conventional standard clinical TMS treatments is challenging. This is attributed to the fact that, for the UNSYNC group, the patients still received TMS pulses adjusted to their personalized prefrontal alpha frequency but with a random phase. Another notable limitation is the absence of a sham intervention. The sensations and auditory aspects associated with TMS may have influenced the quantified acute effects on the fMRI signal. The study’s double-blind design ensured that both groups experienced these factors during the intervention similarly. However, future studies employing sham-controlled interventions are needed to rule out any potential confounding effects caused by the sensory stimulation during TMS. Furthermore, this study focused on optimizing the timing of TMS targeting, but we did not employ a strategy for precise and individualized spatial targeting (EEG F3 electrode was used as a target). Our findings suggest that varying responses across different ROIs near the L-DLPFC may influence the propagation of the effects to distinct areas and, consequently, the different extent of clinical outcome associations. The observed spread of TMS-induced acute effects, potentially following functional connectivity patterns as suggested by our results, underscores the need for future research to optimize spatial targeting of TMS based on individualized functional connectivity and possible electric field modeling^66^. In this EEG-synchronized rTMS treatment, synchronization of TMS pulses with the personalized optimal phase, which is associated with the largest evoked response in the cingulate, proves to be a novel approach for TMS timing target selection. Building on the current study’s results, future investigations could explore optimizing timing selection based on the evoked responses in specific brain networks, such as the cognitive control and limbic networks, or the modulated connectivity between the L-DLPFC and the posterior sgACC in the limbic system. Additionally, employing network neuroscience methodologies to select the best TMS timing could provide novel intervention approaches that might lead to potentially optimized interaction, integration, or segregation across brain networks, potentially leading towards optimized TMS effects on behavior or TRD treatment.

In conclusion, the present study highlights the importance of quantifying TMS-induced acute effects on brain network systems, including evoked response and modulated functional connectivity. These quantifications demonstrate brain areas and circuits engaged in underlying TMS perturbation and potentially their involvement in the therapeutic effects of rTMS treatment in depression. Additionally, the state-dependent analyses established that, by conditioning on different phases of the prefrontal alpha oscillation, the spread of TMS-induced activity follows the functional connectivity of the stimulation site. Finally, in an exploration analysis with limited sample size, longitudinal changes in the state-specific TMS-modulated connectivity in the brain circuit of the stimulation site and sgACC are associated with the clinical outcome. These results suggested that EEG-synchronized rTMS engages specific brain networks (right limbic and bilateral cognitive control networks) and brain circuits (L-DLPFC and right posterior sgACC) toward depression treatment. These results carry significant implications for the treatment of TRD and the development of more precise and personalized TMS treatment protocols, with a focus on both target selection and timing parameters.

## Methods

### Participants and procedure

This randomized and double-blinded clinical trial study was conducted at the Medical University of South Carolina (MUSC) Institute of Psychiatry (ClinicalTrials.gov ID: NCT032421808 ^67^). The experimental procedures of our study and the recruitment process were approved by the MUSC institutional review board. All patients have provided informed consent to participate in the study, and written consent was obtained from the participants.

Thirty-four TRD patients were enrolled in the study, and six subjects were excluded due to voluntary withdrawal, claustrophobia, or inability to complete the pre-treatment MRI scan. Pre-treatment fET data from the remaining twenty-eight patients (mean age ± SD = 45 ± 13 years, female/male = 19/9) were collected. Then, these twenty-eight patients were randomized into the SYNC or UNSYNC group before the treatment, with fifteen patients in the SYNC group and thirteen patients in the UNSYNC group. Ten patients in the SYNC group completed the study, with five patients excluded due to 1) life stress, 2) inability to adhere to the treatment schedule, 3) equipment unavailable, and 4) patients opting not to complete it. Ten patients in the UNSYNC group completed the study, with three patients excluded due to 1) hospitalization, 2) equipment unavailable, and 3) opting not to complete. All enrolled patients had a baseline HRSD (Ham-D 28-item) score greater than or equal to twenty at the time of enrollment. Details of patient enrollment and inclusion/exclusion criteria were described elsewhere^31^.

After enrollment, we performed an integrated fET scan on the patients to determine an individualized optimum phase *φ*_opt_, which was defined as the phase that produced the largest BOLD signal increase in the dACC. During the closed-loop EEG-rTMS treatment, patients in the SYNC group received rTMS treatment with the target phase set as the individualized *φ*_opt_, while for the patients in the UNSYNC group, the target phase was randomly selected from a uniform distribution between 0 to 2π. As the rTMS was delivered at the patients’ daily individualized alpha frequency (6-13 Hz), and the initial pulse was delivered at the pre-defined target phase, patients in the SYNC group received rTMS pulses synchronized to the individualized optimum phase, and patients in the UNSYNC group received rTMS pulses that were unrelated to the patients’ optimum phase (initial pulse was fired randomly). Each patient received one treatment session each weekday for six weeks, with a total of thirty closed-loop EEG-rTMS treatment sessions. During the treatment sessions, the TMS coil was placed at the L-DLPFC (F3 electrode) with the dose of 120% motor threshold, 40 pulses per train, and 75 pulse train per session (3000 pulses in total per session). Details of the closed-loop EEG-rTMS treatment were described in ^31,48^. After the treatment, post-treatment fET scans were acquired from the remaining patients. Each patient’s HRSD was assessed during the pre-and post-treatment fET scans and before the first treatment of each week. The experimental procedure and data analyses are illustrated in Fig. 1.

### Data acquisition and preprocessing

An integrated fET instrument was developed and used in this study^68^, where simultaneous EEG-fMRI data were acquired from the patients while receiving single-pulse TMS at L-DLPFC. The EEG data were acquired with a custom MR-compatible bipolar EEG cap (36 electrodes, sampling rate = 488 Hz, Innovative Technologies, CA, USA). A Siemens 3T Prisma MRI Scanner was used to acquire fMRI data with a custom 12-channel head coil (Rapid MR International, LLC, Columbus, OH, USA). A modified MR-compatible TMS coil was used (MagStim Rapid2) and configured to 100%-120% intensity of each subject’s motor threshold.

Functional MRI data were collected with T2*-weighted multi-echo multiband pulse sequence (CMRR, University of Minnesota) with parameters as follows: TR = 1750 ms with a 200 ms gap, TE1 = 11.20 ms, TE2 = 32.36 ms, TE3 = 53.52 ms, flip angle 59 degrees, multiband acceleration factor = 2, voxel size 3.2 x 3.2 x 3.2 mm, matrix size = 64 x 56 x 38, 233 volumes, six runs. T1-weighted structural image was acquired with a 32-channel Siemens head coil (Multiecho MPRAGE, TR = 2530 ms, TE= 1.55/3.26/5.12/6.98 ms, voxel size 1 x 1 x 1 mm, 176 slices). Images with reversed phase encoding direction were acquired for geometric distortion correction. Single-pulse TMS was delivered to the L-DLPFC (marked under the EEG F3 electrode, Beam F3 locator) at the beginning of the TR gap, with inter-trial intervals drawn from a uniform distribution (4-6 TRs). Details of data acquisition, EEG data preprocessing, and alpha phase estimation were described in^35^.

Functional MRI data were preprocessed with AFNI^69^. Firstly, slice timing correction was performed on each echo time series data. At the same time, susceptibility distortion correction warping fields were estimated from echo-planar imaging (EPI) images with opposite phase encoding direction. The co-registrations between functional and structural images were performed using local Pearson correlation^70^. Additionally, motion correction parameters were estimated from the first echo time series data. Then, the estimated distortion correction warping field, motion parameters, and co-registration transformations were concatenated and applied to the slice timing corrected data of each echo time series. The transformed echoes were combined into a single dataset with the optimally combined weights^71^. Lastly, the combined time series dataset was spatially smoothed with a Gaussian kernel of full width at half maximum (FWHM) 5 mm and scaled to the percent of mean signal level with a mean of 100. Motion-related nuisance signals (six standard head motion parameters and their temporal derivatives) were regressed out from the preprocessed fMRI data before further analysis. No global signal regression was applied. Spatial normalization was performed by using FLIRT^72^ and FNIRT^73^ from the FSL software package, where the structural T1-weighted MR image was initially affine transformed and then non-linearly registered to the MNI152 (the nonlinear 6th generation atlas from FSL) brain image.

Structural T1-weighted MR images were processed with the FreeSurfer pipeline^74^, including brain tissue segmentation and cerebral cortex surface reconstruction. Local-global Schaefer cortical parcellation atlas^75^ was used to define cortical ROI and network systems, which is an fMRI-based parcellation approach integrating local gradient and global similarity approaches based on resting state fMRI data from 1489 young adults. The Schaefer atlas was transformed to each subject’s cortical surface through surface-based registration using FreeSurfer. Then, the ROI surface areas were projected into the volumetric space, where the cerebral cortex was parcellated into 400 ROIs. The Schaefer atlas classified these 400 ROIs into 34 brain networks with 106 network nodes, with each node consisted of adjacent subregions of each node. The L-DLPFC EEG F3 stimulation site ROI was defined in the MNI152 space with a 10 mm radius sphere centered at the coordinate MNI = [-37 26 49]^76^. For each subject, the L-DLPFC EEG F3 ROI was warped into each subject’s space with the estimated spatial normalization parameters and intersected with the gray matter mask. To maintain consistency in brain parcellation, we also assessed L-DLPFC ROIs near the EEG F3 stimulation site by selecting the ROIs from the Schaefer atlas at the nearest distance to the EEG F3 coordinates. Specifically, we included L-DLPFC ROIs near the stimulation site: 1) dorsal-PFC in the subnetwork A of the DMN; 2) lateral-PFC in the subnetwork B of the DMN 3) lateral-PFC in the salience/ventral attention network (subnetwork B).

### Whole-brain general linear model analysis

To investigate the propagation patterns of TMS-induced effects on whole-brain BOLD signal, we performed an event-related GLM analysis, where each TMS pulse was modeled as an instantaneous event, convolved with a set of optimized basis functions known as FMRIB’s linear optimal basis set (FLOBS)^77^. The use of FLOBS compared to the canonical hemodynamic response function (HRF) allows more freedom in the variability of hemodynamic response, as we expect the TMS-induced brain activity might be different from that evoked by conventional fMRI task stimuli, and the FLOBS allows us to control the HRF variabilities across different brain regions. The combined parameters from FLOBS were carried out for the group-level analysis. Specifically, we computed the signed root mean square of the regression parameter estimates from the subject-level^78^. As the distribution of the combined statistic is non-Gaussian, in the group-level analysis, we used FSL ‘Randomise tool’^49^ for a nonparametric permutation statistical testing (10000 permutations; p < 0.05 with multiple comparison correction across space using voxel-wise family-wise error).

### Quantification of TMS-evoked BOLD response

To assess subject-wise TMS-induced acute effects and quantify its inter-subject variability, we quantified both the spatial extent and amplitude of TMS-evoked BOLD response across brain networks. We defined the spatial extent of the induced response as the percent coverage of each network by the propagation pattern of TMS-evoked BOLD response. Specifically, based on the subject-wise whole-brain GLM analysis results, the spatial extent of each network activated by the TMS was computed as the number of significantly activated voxels divided by the total number of voxels within each network. Then, to quantify the amplitude of TMS-evoked BOLD response for each brain network, we performed ROI analysis, where the mean BOLD signal of each ROI was modeled by the TMS pulse timing regressor (TMS pulse timing boxcar function convolved with the FLOBS basis set) with GLM. The signed root mean square of the regression parameter estimates was summed across ROIs within each network to compute the amplitude of the network’s induced response.

### Functional connectivity and hub analysis

To examine the TMS-induced acute modulation of FC, we performed a whole-brain PPI analysis^51–53^. Firstly, the BOLD signal from each parcellated cortical region was extracted with SPM^79^. Then, to compute the TMS-dependent functional connectivity, we used multiple regression, where the BOLD signal of an ROI *A* was modeled as a linear combination of multiple independent variables including 1) BOLD signal of ROI *B*; 2) TMS-induced BOLD response (a boxcar function representing the timing of TMS pulses convolved with FLOBS); 3) PPI interaction between the BOLD signal at ROI *B* and the timing of TMS pulses; 4) nuisance variables (motion parameters, large motion volumes, BOLD signal in white matter, and BOLD signal in lateral ventricle). The interaction between the BOLD signal from one ROI and the task regressor was modeled by first deconvolving the BOLD signal from the canonical HRF and then being multiplied by the task timing boxcar function, and lastly re-convolving with the HRF. As the PPI analysis focuses on the second-order task modulation, controlling the first-order task modulation (task-evoked mean activation) is necessary, as the task-evoked mean activation could be a potential confounder and drives the false positive observed interaction between regions^80^. Specifically, FLOBS-based task regression was performed to account for the task-evoked mean activation, which has been shown to achieve relatively low false positive results in controlling the confounds^80^. The TMS-modulated functional connectivity matrix (beta estimates of the interaction term in the GLM model) between each pair of ROI A and *B* was extracted, symmetrized (averaging the upper and lower triangles), and summarized within/between parcellated brain networks. Finally, we assessed positive/negative node strength by computing the sum of all positive/negative connection weights associated with each node.

In addition to the TMS-modulated connectivity, we also assessed correlation-based functional connectivity. The seed-based functional connectivity analysis of the L-DLPFC was performed with a mixed-effects model. The functional connectivity of each patient was computed based on the Pearson correlation between the time series of each ROI and the time series of each voxel in the brain. Before computing the Pearson correlation, the TMS-related variabilities (modeled with FLOBS basis sets) were regressed out from the preprocessed fMRI time series. Then, each subject’s functional connectivity map was transformed into a z-score with Fisher’s Z transformation and threshold at p < 0.01. One sample student’s t-test was performed at the group level to obtain the seed-based functional connectivity t-value map of the ROI.

### Brain-state dependency analysis

The aim of this analysis was to assess whether single-pulse TMS delivered at different timing relative to the prefrontal alpha oscillation modulates brain network systems differently. To examine the brain-state dependency effect of TMS delivery, we used the phase of prefrontal alpha oscillation to index the brain state. Specifically, we extracted prefrontal alpha oscillation from the EEG signals at channels FP1, F3, and F7, and the alpha phase at TMS onset was estimated (details in^35^). Based on the alpha phase, TMS trials were grouped into four bins (bin 1: −π to −1/2 π, bin 2: −1/2 π to 0, bin 3: 0 to 1/2 π, and bin 4: 1/2 π to π), and we investigated the TMS effects on brain network systems for the trials in different phase bins.

Firstly, to examine the brain-state dependency of the TMS evoked response, we used GLM to model the BOLD signal at L-DLPFC (EEG F3 stimulation site), with the trials in each phase bin as a separate regressor (Fig. 6). This analysis aimed to identify two TMS trial conditions for each subject: 1) HLP condition was defined as the condition where a high TMS evoked response was introduced at the stimulation site (L-DLPFC); 2) LLP condition was defined with a low evoked response at L-DLPFC. Because the phase bins groups differ in the number of trials and the temporal spacing, this might potentially lead to biased estimation of BOLD response and the trial conditions identification. To prevent such bias during HLP/LLP conditions identification, we performed bootstrapping where same number of trials were randomly chosen for each phase bin to construct the regressors. Here, we set the selected trial number as eighty percent of the trial number in the phase bin with the smallest number of trials, and we repeated the bootstrapping process with 500 iterations. For each subject, the phase bins that generated the highest and lowest evoked response at L-DLPFC were identified as the subject-wise HLP and LLP bins, respectively. Then, we assessed the contrast between the conditions of HLP and LLP bins. Whole-brain GLM analysis was performed to identify other brain areas with a significantly higher evoked response for the TMS trials in the HLP bins compared to that in the LLP bins (correlates of the HLP-vs-LLP contrast). The beta-weight maps of the estimated contrast were carried out to the group level, and a voxel-wise t-test against zero was performed with uncorrected threshold of p < 0.001.

**Fig. 6.**
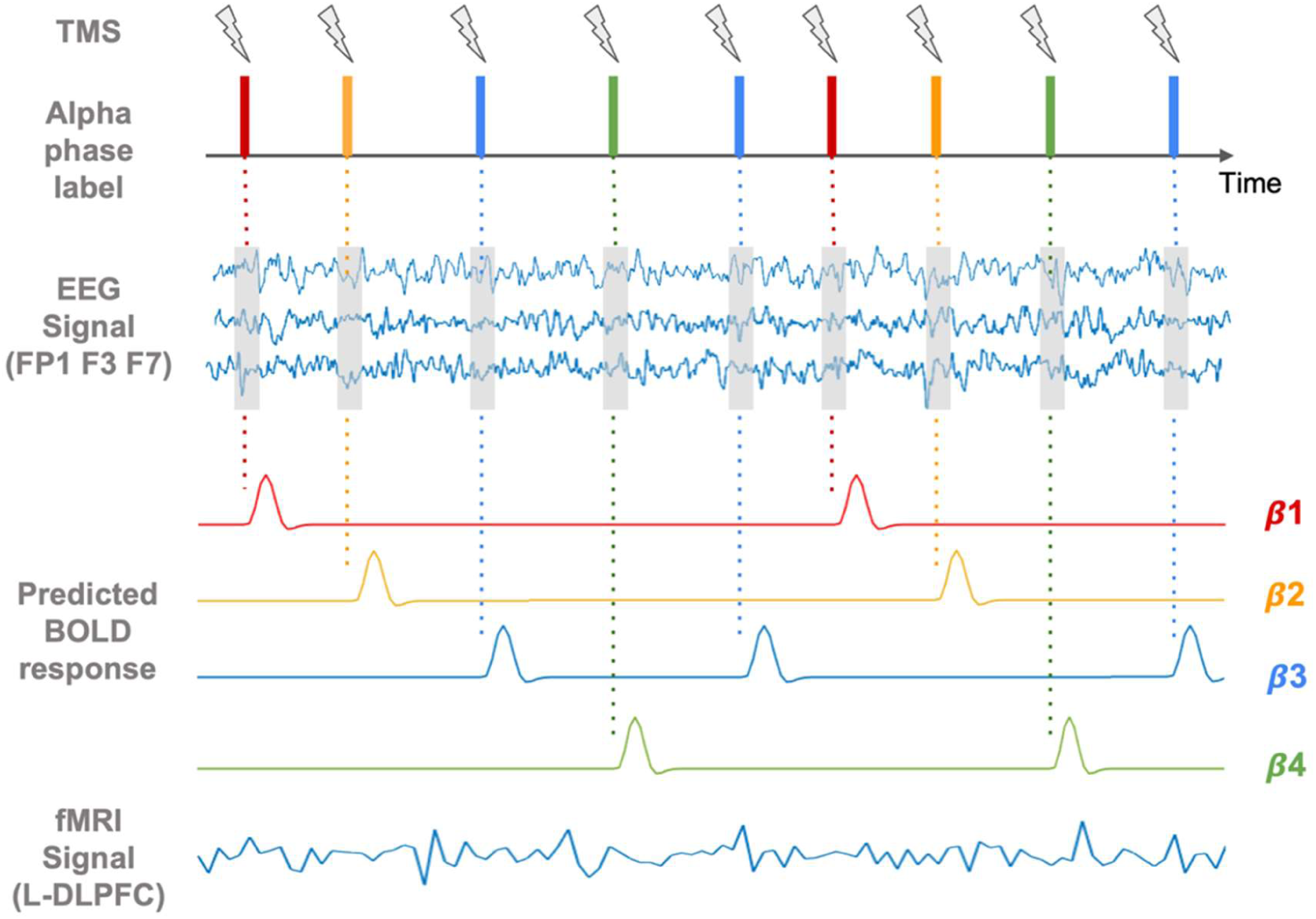
Brain-state dependency analysis of TMS-induced acute effects using alpha phase from the prefrontal EEG signal. TMS trials were grouped into four categories with different prefrontal alpha phase bins (illustrated with different colors), where the prefrontal alpha oscillation was extracted from the concurrent EEG recordings at electrodes FP1, F3, and F7. Effects from the TMS trials in different phase bins were modeled with general linear modeling, where the BOLD signal at L-DLPFC (EEG F3 stimulation site) was modeled with the TMS trials in each phase bin as a separate regressor. The phase bins that generated the highest and lowest BOLD response at L-DLPFC were identified as the subject-wise high-load-phase (HLP) and low-load-phase (LLP) conditions, respectively.

Additionally, we also examined the brain-state dependency of TMS-modulated functional connectivity. Similar to the PPI analysis in the previous section, the same approach was adapted to assess the modulated FC for the TMS trials in each phase bin. In this analysis, the PPI interaction terms include four regressors, and each of them models the interaction between the BOLD signal at ROI *B* and the timing of TMS pulses in each phase bin.

### Associations between TMS effects and clinical response

Pearson correlation coefficient was used to assess the relationship between neuroimaging measurements of TMS effects and the clinical response. The clinical response was quantified as the percent improvement of the HRSD score since the baseline, i.e. 100 × (pre-treatment score – post-treatment score)/pre-treatment score. We also assessed whether the neuroimaging measurements (TMS-induced response and TMS-modulated functional connectivity) at baseline pre-treatment scan are predictive biomarkers of the clinical response. Additionally, to explore the potential mechanisms of action underlying EEG-synchronized rTMS treatment, we also examined the longitudinal changes in the neuroimaging measurements of TMS effects. Specifically, we quantified the changes in the TMS-induced response, modulated functional connectivity by all TMS trials, and modulated functional connectivity by TMS trials in the HLP bin (state-specific modulation). Finally, we tested their associations with the clinical response as well.

## Data Availability

All data produced in the present study are available upon reasonable request to the authors.

## Data and code availability

The datasets analyzed in the current study are available from the corresponding author on reasonable request. Custom codes used in this work are available at https://github.com/hehengda/fMRI_EEG_TMS.git.

## Acknowledgments

This work was supported by the National Institute of Mental Health (MH106775), a Vannevar Bush Faculty Fellowship from the US Department of Defense (N00014-20-1-2027) a Center of Excellence grant from the Air Force Office of Scientific Research (FA9550-22-1-0337) and by DARPA (HR00112320032).

## Author contributions

Design of research: H.H., T.R.B., M.S.G., and P.S.; Methodology: H.H., X.S., J.D., J.F., R.I.G., T.R.B., M.S.G., and P.S.; Formal analysis: H.H.; Data collection: J.D., J.F., G.T.S., S.H., R.I.G., T.R.B., M.S.G., and P.S.; Writing original draft: H.H.; Writing, review, and editing: H.H., X.S., J.D., J.F., J.R.M., G.T.S., S.H., L.H., S.P.P., H.Y., L.M.M., R.I.G., T.R.B., M.S.G., and P.S.; Funding and supervision: L.M.M., R.I.G., T.R.B., M.S.G. and P.S.

