## Supplementary Information for "TMS-induced modulation of brain networks and its associations to rTMS treatment for depression: a concurrent fMRI-EEG-TMS study"

[1]Department of Biomedical Engineering, Columbia University, New York, 10027, NY, USA.
[2]Center for Biomedical Imaging, Medical University of South Carolina, Charleston, 29425, SC, USA.
[3]Department of Orthopedic Surgery, Columbia University Irving Medical Center, New York, 10032, NY, USA.
[4]Department of Radiology and Radiological Science, Medical University of South Carolina, Charleston, 29425, SC, USA.
[5]Department of Neurology, University of Chicago, Chicago, 60637, IL, USA.
[6]Department of Psychiatry and Behavioral Sciences, Medical University of South Carolina, Charleston, 29425, SC, USA.
[7]Department of Psychiatry, Columbia University Irving Medical Center, New York, 10032, NY, USA.
[8]Stephenson School of Biomedical Engineering, The University of Oklahoma, Norman, 73019, OK, USA.
[9]Center for Healthy Minds, University of Wisconsin-Madison, Madison, 53705, WI, USA.
[10]Ralph H. Johnson VA Medical Center, Charleston, 29401, SC, USA.
[11]Department of Electrical Engineering, Columbia University, New York, 10027, NY, USA.
[12]Department of Radiology, Columbia University Irving Medical Center, New York, 10032, NY, USA.
[13]Data Science Institute, Columbia University, New York, 10027, NY, USA.

- **Supplementary text**

### State-dependency of TMS-induced acute effects and modulation

When examining the TMS modulations on the functional connectivity with PPI analysis, we observed state-specific effects for the high-load phase bins of the L-DLPFC stimulation site. Our results showed significant (FDR-corrected $p < 0.05$) negative modulations on the functional connectivity only for TMS trials in the high-load phase bins between: 1) default mode network (subnetwork A, RH) and default mode network (subnetwork B, RH); 2) default mode network (subnetwork A, RH) and control network (subnetwork A, RH); 3) default mode network (subnetwork A, RH) and control network (subnetwork B, RH). However, when we performed the same analyses for the high-load phase bins of the other L-DLPFC ROIs, no such significant modulations were found after multiple comparison correction.

Additionally, we further examined these network connections. Our results showed that these state-specific modulations were evoked to a different extent based on different L-DLPFC ROIs. For example, the connection between default mode network (subnetwork A, RH) and default mode network (subnetwork B, RH) was strongly modulated by the TMS trials with the high-load phase bin of L-DLPFC EEG F3 stimulation site ($z = -3.8987$, $p < 9.6695e-5$), followed by the non-significant modulations of TMS trials associated with L-LPFC in DefaultB network ($z = -2.9956$, $p < 0.0027$), L-DPFC in DefaultA network ($z = -2.1664$, $p < 0.0303$), and L-LPFC in salience/ventral attention network (subnetwork B, LH) network ($z = -1.8048$, $p > 0.0711$). Likewise, the connection between default mode network (subnetwork A, RH) and control network (subnetwork A, RH) was strongly modulated by the TMS trials with the high-load phase on L-DLPFC EEG F3 stimulation site ($z = -3.9123$, $p < 9.1409e-5$), followed by the non-significant modulations of TMS trials associated with L-LPFC in DefaultB network ($z = -3.3223$, $p < 0.0009$), L-DPFC in DefaultA network ($z = -2.9969$, $p < 0.0027$), and L-LPFC in salience/ventral attention network (subnetwork B, LH) network ($z = -1.6409$, $p > 0.1008$). Lastly, the connection between default mode network (subnetwork A, RH) and control network (subnetwork B, RH) was strongly modulated by the TMS trials with the high-load phase on L-DLPFC EEG F3 stimulation site ($z = -3.7138$, $p < 0.0002$), followed by the non-significant modulations of TMS trials associated with L-LPFC in DefaultB network ($z = -2.6845$, $p < 0.0073$), L-DPFC in DefaultA network ($z = -1.7967$, $p > 0.0723$), and L-LPFC in salience/ventral attention network (subnetwork B, LH) network ($z = -1.6925$, $p > 0.0905$). These results suggest the spatial specificity of the L-DLPFC locations when examining the TMS state-specific effects, indicating that when the TMS trials induce a different extent of evoked responses at the L-DLPFC locations, the spatial specificity near the L-DLPFC is related to different extent of evoked modulations on the connections between these brain circuits.

- **Supplementary figures**

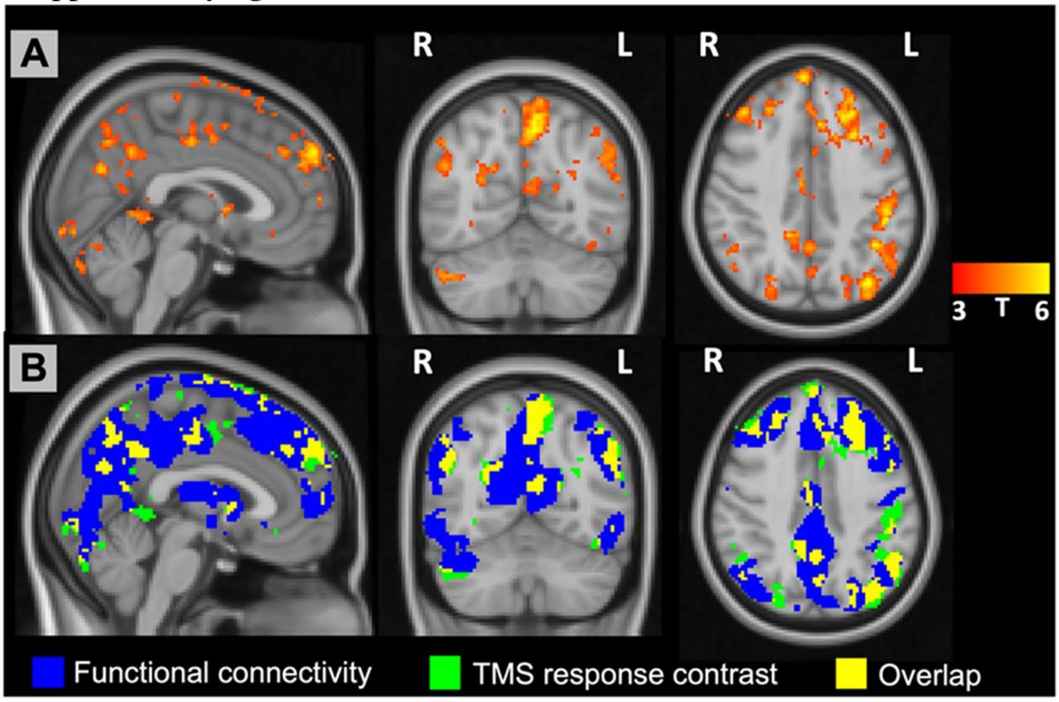


Figure S1. State-dependency analysis of TMS-evoked BOLD response (LH-dorsal-PFC in the default mode network (subnetwork A, LH)). (A) TMS response contrast between the conditions of TMS trials in the high-load-phase bins and low-load-phase bins (t-value; uncorrected $p < 0.001$). These regions have a stronger BOLD response to TMS perturbation when TMS induced a stronger BOLD response at the L-DLPFC ROI (LH-dorsal-PFC in the default mode network (subnetwork A, LH)). (B) Spatial overlap between the TMS response contrast and L-DLPFC seed-based functional connectivity.

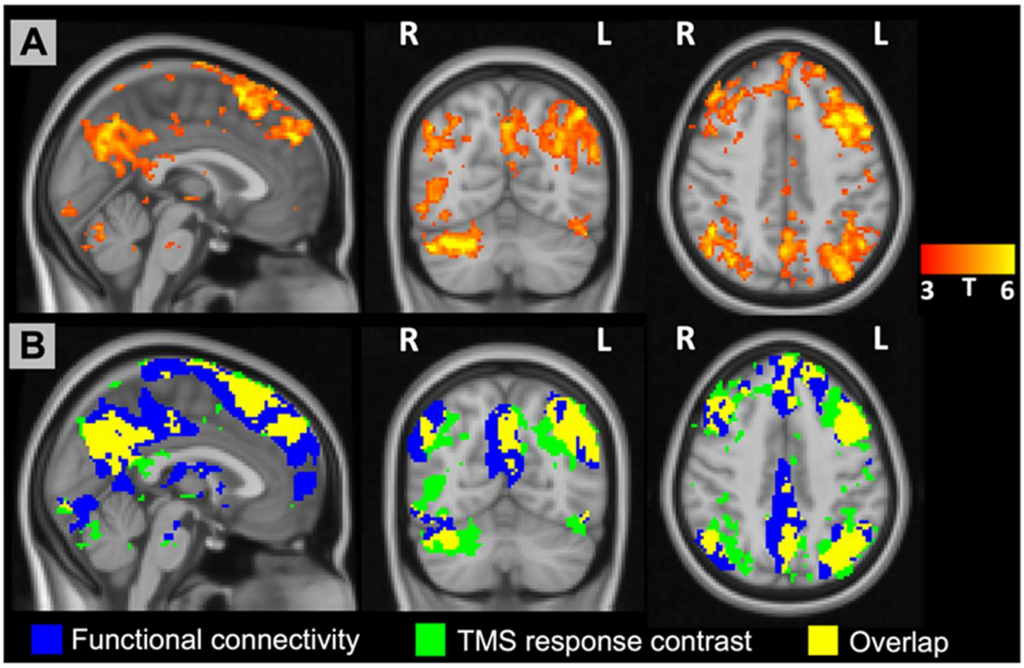


Figure S2. State-dependency analysis of TMS-evoked BOLD response (LH-lateral-PFC in the LH-DefaultB network). (A) TMS response contrast between the conditions of TMS trials in the high-load-phase bins and low-load-phase bins (t-value; uncorrected $p < 0.001$). These regions have a stronger BOLD response to TMS perturbation when TMS induced a stronger BOLD response at the L-DLPFC ROI (LH-lateral-PFC in the LH-DefaultB network). (B) Spatial overlap between the TMS response contrast and L-DLPFC seed-based functional connectivity.

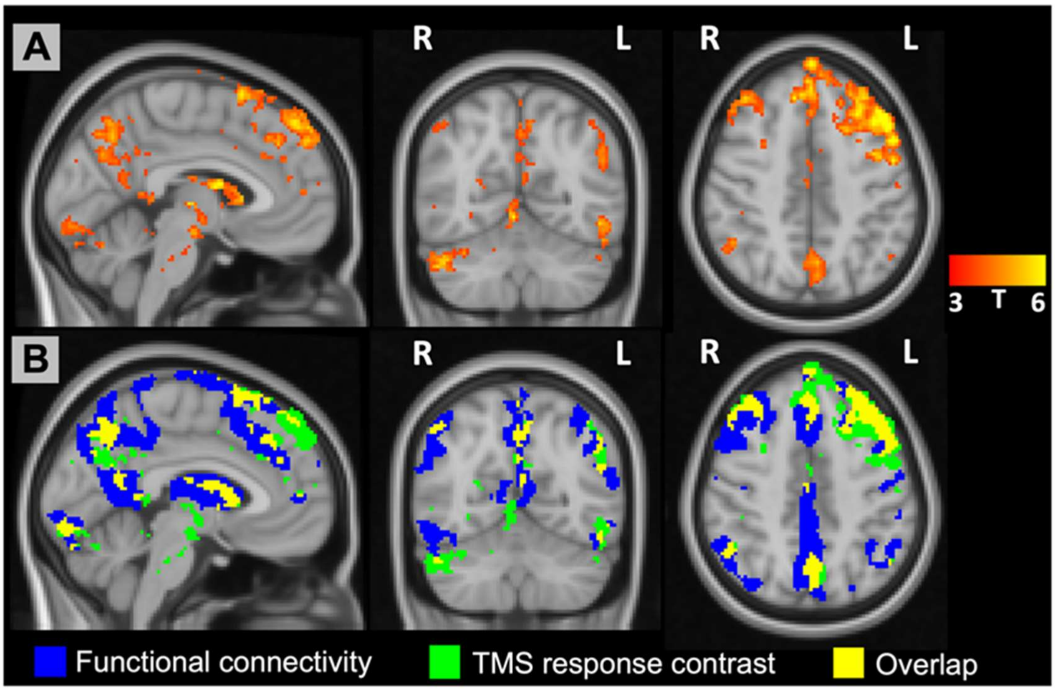

Figure S3. State-dependency analysis of TMS-evoked BOLD response (LH-lateral-PFC in the LH-SalVentAttnB network). (A) TMS response contrast between the conditions of TMS trials in the high-load-phase bins and low-load-phase bins (t-value; uncorrected $p < 0.001$). These regions have a stronger BOLD response to TMS perturbation when TMS induced a stronger BOLD response at the L-DLPFC ROI (LH-lateral-PFC in the LH-SalVentAttnB network). (B) Spatial overlap between the TMS response contrast and L-DLPFC seed-based functional connectivity.
